# Reaching the 100 by 2027 target for universal access to rapid diagnostic tests for tuberculosis in Africa: in-sight but out of reach

**DOI:** 10.1101/2025.08.26.25334429

**Authors:** Lucy Mupfumi, Tapson Nyondo, Judith Mzyece, Vincent Kampira, Moussa Condé, Aloni L. Muriel, Manga H.R. Arsène, Joselyne Ndayihimbaze, Charles Lamou Ki-zerbo, Jean Njab, Caroline Bih, Mariamo Ibraimo Assane, Elishebah Mutegi, Michael Maina, Francis Ocen, Silver Mashate, Collins Otieno Odhiambo

## Abstract

Although WHO recommended rapid molecular diagnostic tools for TB over a decade ago, only about one-third of diagnostic sites in high burden countries have access to them. We sought to understand the barriers and facilitators to reaching universal access to WHO-recommended rapid diagnostics (WRDs) in a selection of African countries. Data for 24 African countries were sourced from the WHO Global TB database to evaluate the proportion of notified people with TB tested with a WRD between 2021 and 2023, and to assess access to WRDs as defined in the WHO Diagnostic Standard. Additionally, TB program staff from six countries were surveyed to identify key factors influencing the implementation of WRD-based diagnostic algorithms. Across the 24 countries, 61% of people with TB were tested with a WRD in 2023. Only in 7 countries was this proportion above 80%, with 8 countries reporting below the global average of 48%. This proportion increased by just 6% between 2021 and 2023, with downward trends observed in 7 countries. Predominant health system barriers included supply chain disruptions, inadequate funding, staffing shortages and weak sample transportation systems. Facilitators included policy and strategic planning, expansion and optimisation of molecular diagnostic capacity, and integrated sample transport networks. Only 7 of the 24 African countries analysed are on track to meet the target for universal access to WRDs by 2027. Persisting health system barriers pose a threat to diagnostic networks and limit the effective use of WRDs. Strategic planning to address these barriers is needed to optimise the use of WRDs and maximise their impact on the TB care cascade.

## Introduction

Over a decade has passed since WHO first issued the recommendation on the use of Xpert MTB/RIF for the rapid detection of TB and rifampicin resistance. More recently, new classes of nucleic acid tests that can be placed at different tiers of the health system and capable of detecting isoniazid and fluoroquinolone resistance are now recommended (1). Yet, diagnostics remain the weakest link in the TB care cascade (2), with close to 3 million people with TB neither diagnosed nor treated annually (3). The slow pace of adoption of WHO-recommended rapid diagnostics (WRDs) contributes significantly to this persisting diagnostic gap. Only 48% of individuals with new or relapse TB were tested with a WRD as the initial test in 2023 (3), suggesting that diagnosis still relies on smear microscopy, a test with inferior sensitivity compared to WRDs. This also contributes to the gap in drug resistance detection, with only half of the estimated 400,000 people with drug-resistant TB diagnosed each year.

The 2018 United Nations High-Level Meeting (UNHLM) on TB outlined bold commitments by heads of states and governments to ending the tuberculosis epidemic globally by 2030 in line with the 2030 Agenda for Sustainable Development (4). However, by 2023, many of these targets were largely unmet, with the biggest shortfalls in the funding target (5). Recognising the need to revitalise commitments and actions to accelerate efforts to End TB, a second UNHLM on TB was held in 2023 to set targets for the next 5 years. These included a new target to ensure universal access to WRD tests for all people with TB by 2027 (6).

Unfortunately, the slow pace of uptake of rapid molecular diagnostics risks the targets remaining aspirational. Despite a diversified landscape of WRDs, less than a third of the diagnostic units in high burden TB countries have access to WRDs (7). Therefore, in 2023, WHO issued the WHO Standard: Universal Access to Rapid TB Diagnostics specifying twelve benchmarks to be tracked across the diagnostic cascade (7).

To understand progress towards universal access to WRDs aligned to the WHO diagnostic standard, we analysed data submitted to the WHO from 24 countries that are part of the Gates Foundation funded Laboratory Systems Strengthening Community of Practice (LabCoP) of the African Society for Laboratory Medicine (ASLM). Furthermore, we surveyed six countries to obtain insights into the challenges and enabling factors for increasing WRD use.

## Methods

### Data review

Data for 24 African countries were extracted from the WHO TB notification, TB laboratories, and TB policies databases (8). Data extracted covered the period 2021 to 2023. We selected the following benchmarks (BM) of the WHO Standard on universal access to rapid tuberculosis diagnostics for this analysis: districts with an algorithm specifying the use of WRDs (BM3), primary health care facilities with access to a WRD (BM4), notified TB tested with a WRD (BM5), WRD testing capacity (BM6), and presumptive TB tested with a WRD (BM8). We calculated the proportions using the specified numerator and denominator from the standard. We defined the following categories to assess progress towards the UNHLM WRD target: Achieved target (90 - 100%), On track (80 - 89%), Making progress (60 – 79%), and Off track (<60%).

### Country stakeholder survey

We sent out a short electronic questionnaire to national TB program staff consisting of diagnostics officers or laboratory focal persons at the central level and the heads of the national TB reference laboratory in six countries (Burundi, Cameroon, Democratic Republic of Congo, Guinea, Zambia and Zimbabwe) to gather insights on barriers and facilitators for WRD use and identified broad themes from the open-ended survey responses. These countries were purposively selected to ensure perspectives reflected diverse experiences across geographical regions. We followed up by e-mail or telephone to probe further selected responses.

### Statistical analysis

Data analysis and visualisation was conducted in R Studio (v2024.12.1+563, Posit Software, PBC). Proportions were compared using the Kruskal-Wallis test. The first author used topic guides that reflected content areas such as barriers and contextual factors influencing WRD use and the themes that emerged during the review of the survey responses to manually organise the data into thematic codes. These were then used to summarise the data by theme.

### Ethics approval

This analysis used programme and publicly available deidentified data that could not be traced back to individual patients. Furthermore, the responses shared by the country stakeholders did not identify individual patient data. Therefore, ethical approval was not required in accordance with local and international guidelines for secondary data analysis.

## Results

We analysed data from 24 African countries; 5 in Central Africa, 6 in Eastern Africa, 7 in Southern Africa and 6 in West Africa. Data on the number of individuals with new or relapse TB who received a WRD as the initial test in 2021 and 2022 was missing for 5 countries. The proportion of people with TB tested with a WRD increased by just 6% from 55% in 2021 to 61% in 2023. Only 7 (29%) of 24 countries reported using a WRD as the initial test for at least 80% of people diagnosed with new or relapse TB in 2023 (Figure 1). However, in eight countries (33%), the proportion of notified people with TB tested with a WRD was below the global average of 48%. There was substantial heterogeneity across regions, with a higher median proportion of notified cases tested with a WRD in Southern Africa (80%, Q1; Q3 62.5, 94.2, p<0.01) compared to the other three regions with a median proportion ranging from 43% (Q1; Q3 21.2, 65.2) in Central Africa to 58% (Q1; Q3 34.3, 68.1) in Western Africa over the 3-year period (Supplementary File 1).

**Fig 1:**
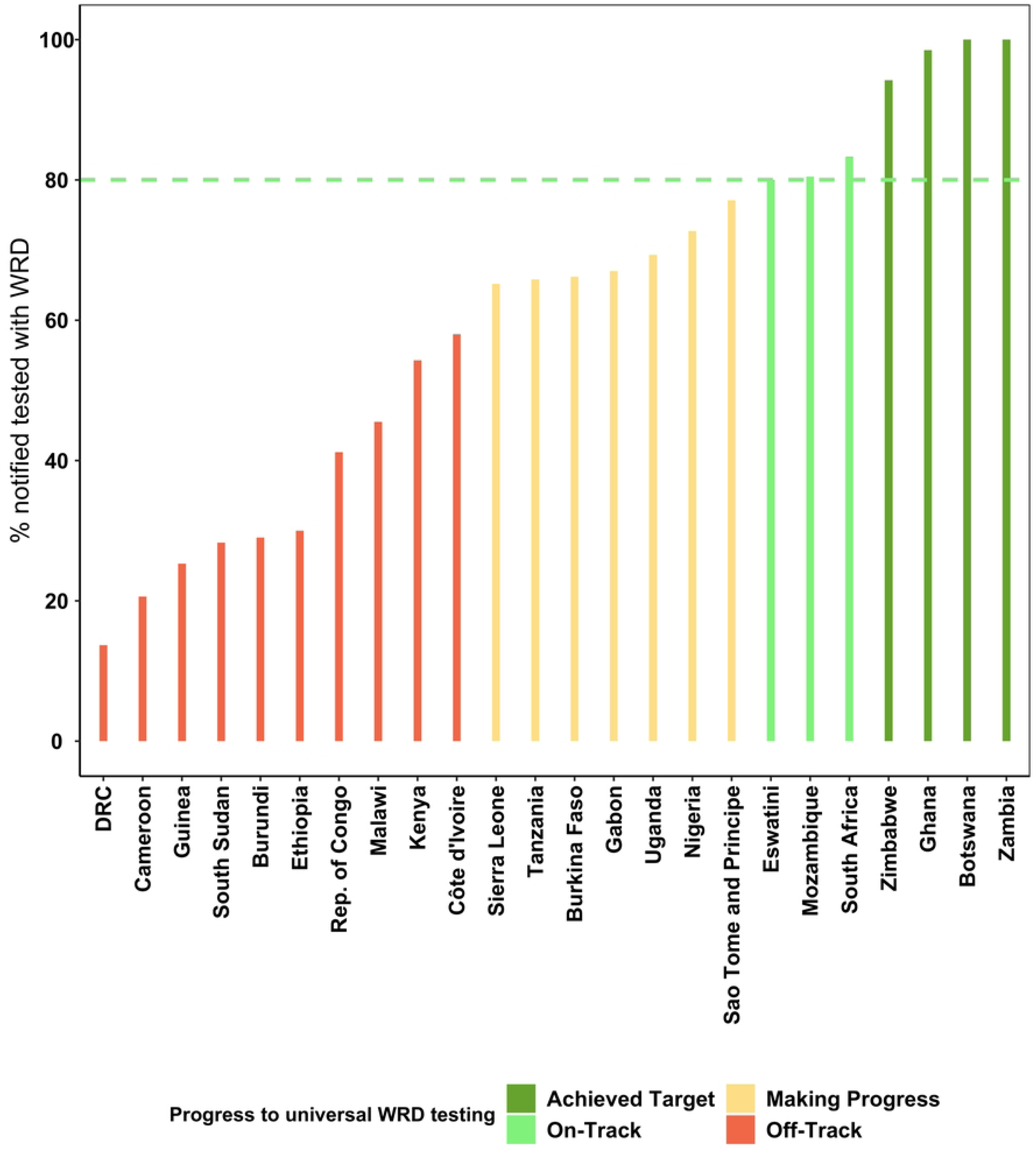
Proportion of people diagnosed with new or relapse TB in 2023 who were tested using a WHO-recommended diagnostic as the initial test across 24 countries. We defined the following categories to show progress towards universal WRD use: Achieved Target (90 - 100%), On-Track (80 - 89%), Making Progress (60 – 79%), and Off-Track (<60%). The green dotted line represents 80% of notified cases tested with a WRD, which we defined as the threshold for “On-Track” to meet the UNHLM target.

When we examined the trend over the years for each country, we observed inconsistencies in the use of WRDs, with a downward trend between 2022 and 2023 observed in 7 countries, 3 of these in the Central African region (Figure 2).

**Fig 2:**
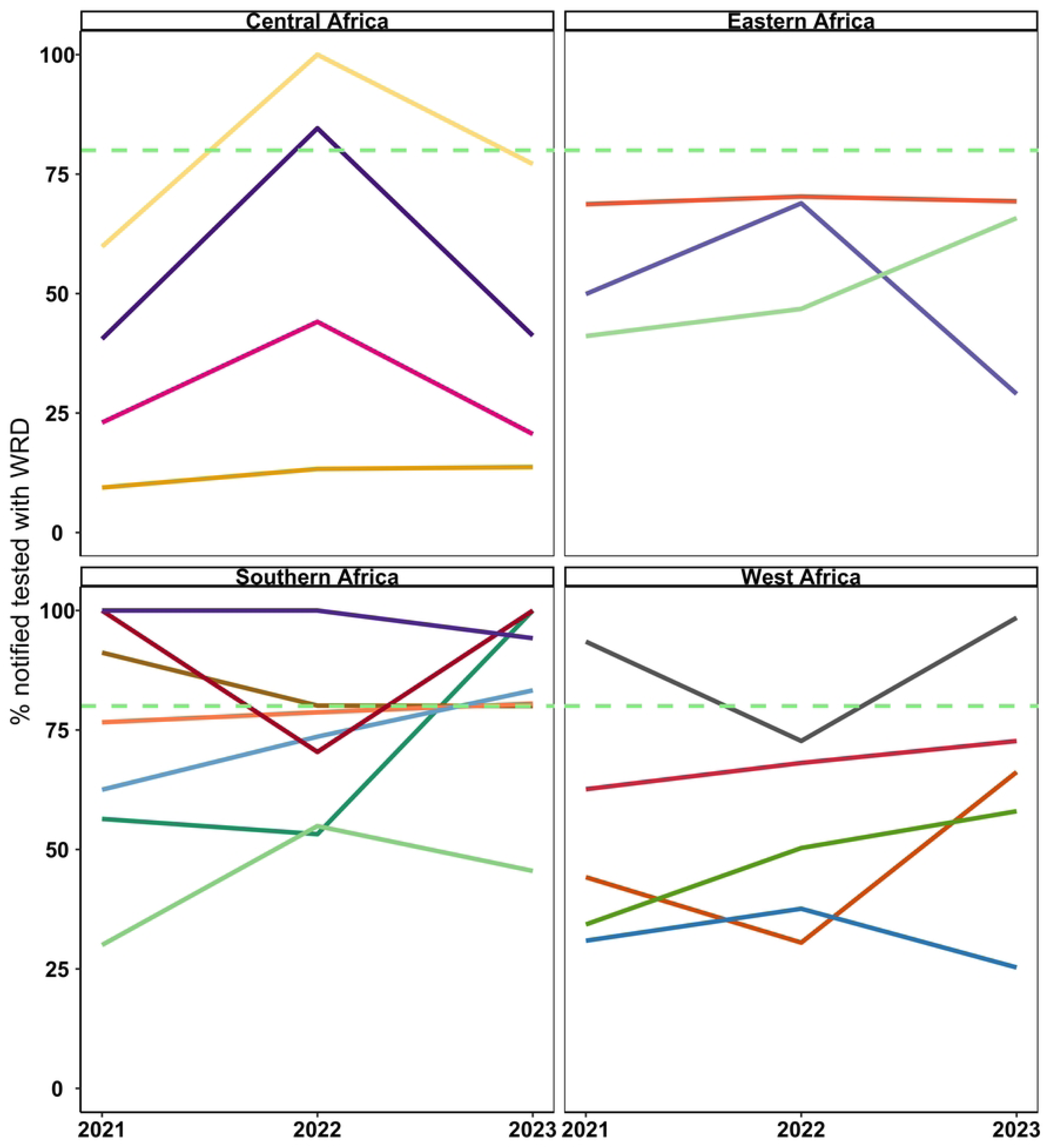
Trend in proportion of people diagnosed with new or relapse TB between 2021 and 2023 who were tested using a WHO-recommended test as the initial diagnostic. The graph shows 19 countries in the ASLM LabCoP network with data available for all three years (Five countries did not have data for 2021 and/or 2022). Line colours represent each country. The green dotted line represents 80% of notified cases tested with a WRD, which we defined as the threshold for “On-Track” to meet the UNHLM target.

Next, we analysed gaps in the TB diagnostic cascade focusing on benchmarks 3,4,5,6 and 8 that address access to and use of WRDs for people with presumptive TB. Thirteen of the twenty-four countries had data for the benchmarks of interest. We observed several inconsistencies in the data. For example, 6 out of the 13 countries (46%) reported having enough test capacity for all people with presumptive TB, yet only 1 country (8%) reported testing at least 90% of the presumptive TB with a WRD. Similarly, 6 countries (46%) reported that all facilities in at least 90% of districts were using algorithms stating the use of a WRD as the initial test, yet only 3 countries (23%) reported testing at least 80% of notified cases with a WRD (Figure 3).

**Fig 3:**
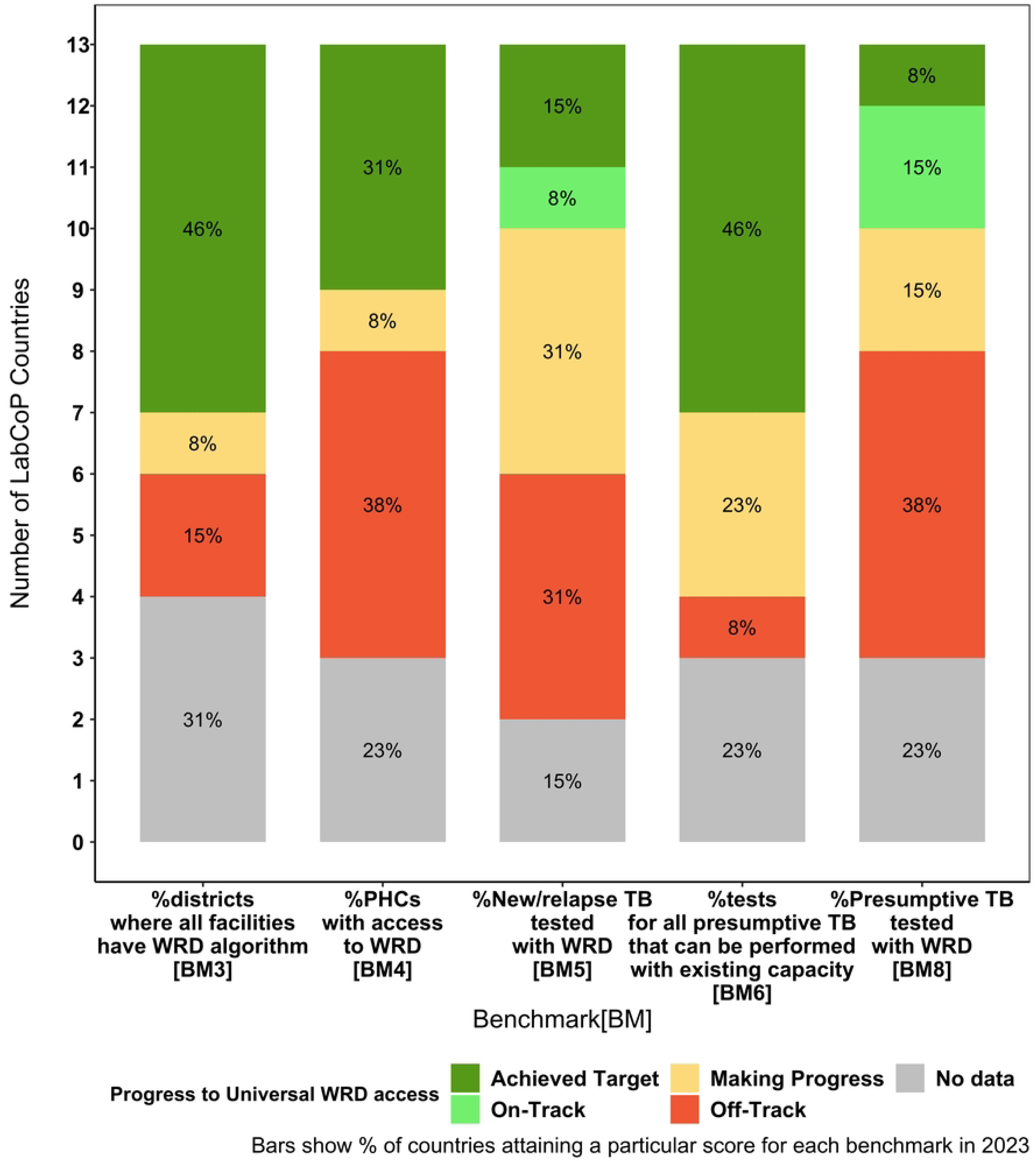
Progress towards meeting the targets of the WHO Diagnostic Standard for 13 LabCoP countries. Bars show percentage of LabCoP countries attaining a particular score for each of the benchmarks defined as: Achieved Target (90 - 100%), On-Track (80 - 89%), Making Progress (60 – 79%), and Off track (<60%).

Furthermore, challenges with reporting data for the benchmarks were evident, as at least 3 countries did not provide either the numerator, denominator, or both for the benchmarks.

When we explored reasons for these trends, three key themes emerged as barriers and facilitators across each step of the diagnostic cascade in the countries surveyed (Table 1). Respondents comprised a TB Diagnostics Officer, a TB Laboratories Coordinator and a Laboratory Focal Point from the National TB Program (NTP), as well as three Managers/Chief/Head of the National TB Reference Laboratory (NTRL).

**Table 1:** Barriers and facilitators for WRD use in six LabCoP countries.

| Theme | Facilitators | Barriers |
| --- | --- | --- |
| 1. Optimise<br>WRD<br>capacity | - National policies and strategies<br>- Program commitments to deploy more molecular tools<br>- Integrated testing<br>- Diagnostic network optimisation (DNO)<br>- Optimise equipment placement for access not just utilisation<br>- Diversify molecular tools | - Stockouts of WRD commodities and reagents<br>- Equipment maintenance |
| 2. Strengthen<br>sample<br>transport &<br>access to<br>testing | - Integrated sample transportation<br>- Awareness and community screening campaigns<br>- Expansion of screening centres | - Poor road networks<br>- Fuel shortages / No fuel allocation<br>- Difficulty accessing isolated areas |
|  |  | - Security concerns in some regions |
| 3. Strengthen financing, staffing & infrastructure | - Laboratory involvement in planning for funding | - Limited funding for rapid diagnostic test devices, reagents or equipment maintenance<br>- Insufficient budget for sample transportation, staffing, training or supervision<br>- Lack of funding for M&E<br>- Insufficient infrastructure for testing, especially at peripheral levels<br>- Low staff motivation |

We then distilled the barriers into five categories and developed mitigation strategies (Table 3) to improve the use of WRDs across all 24 LabCoP countries.

**Table 2:**
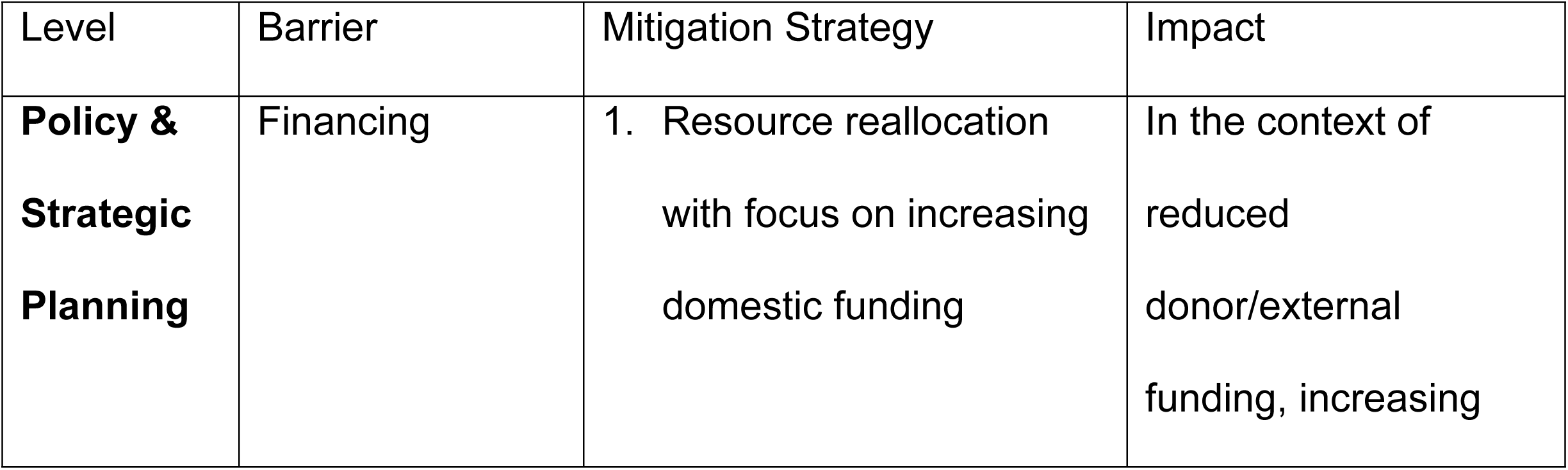
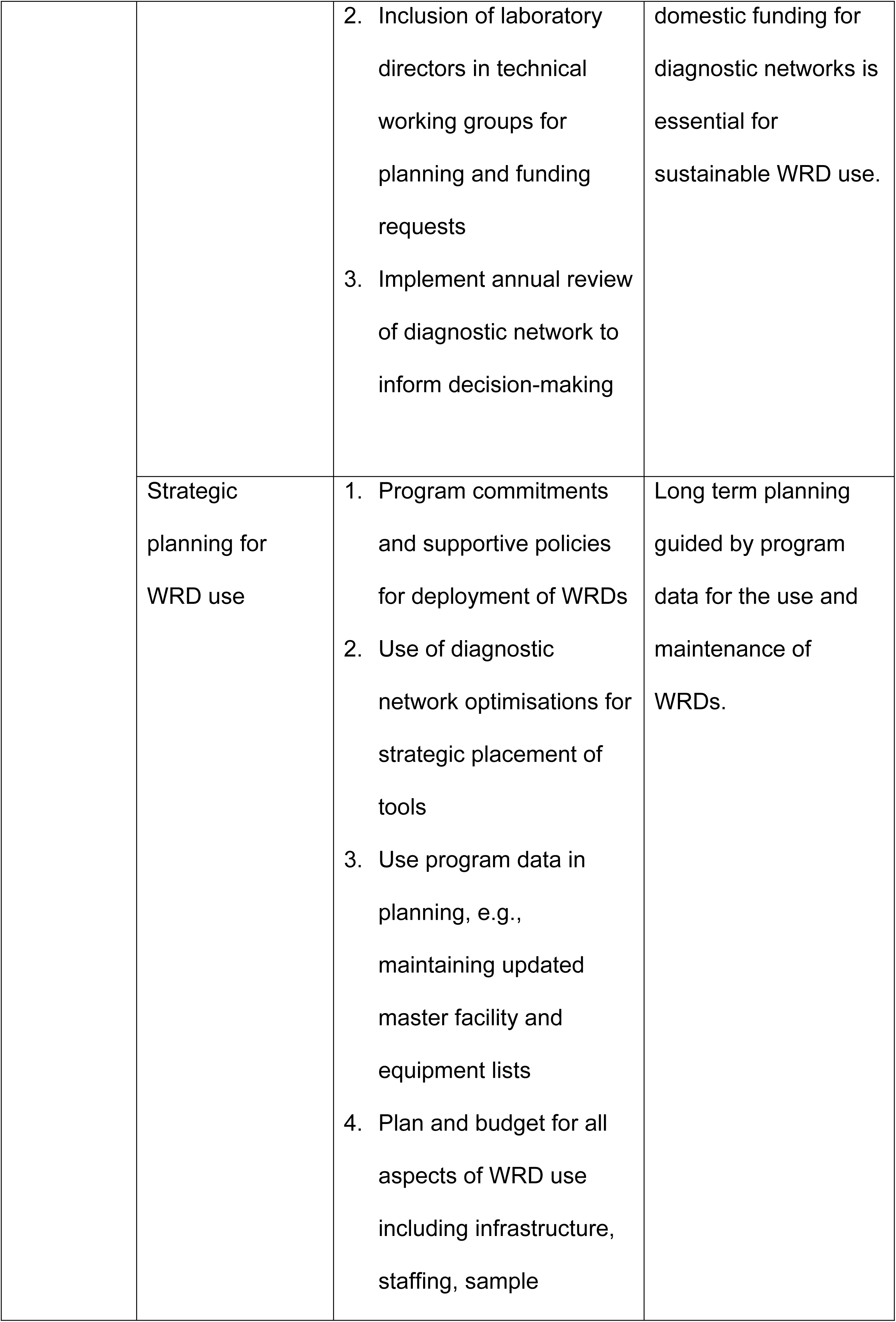
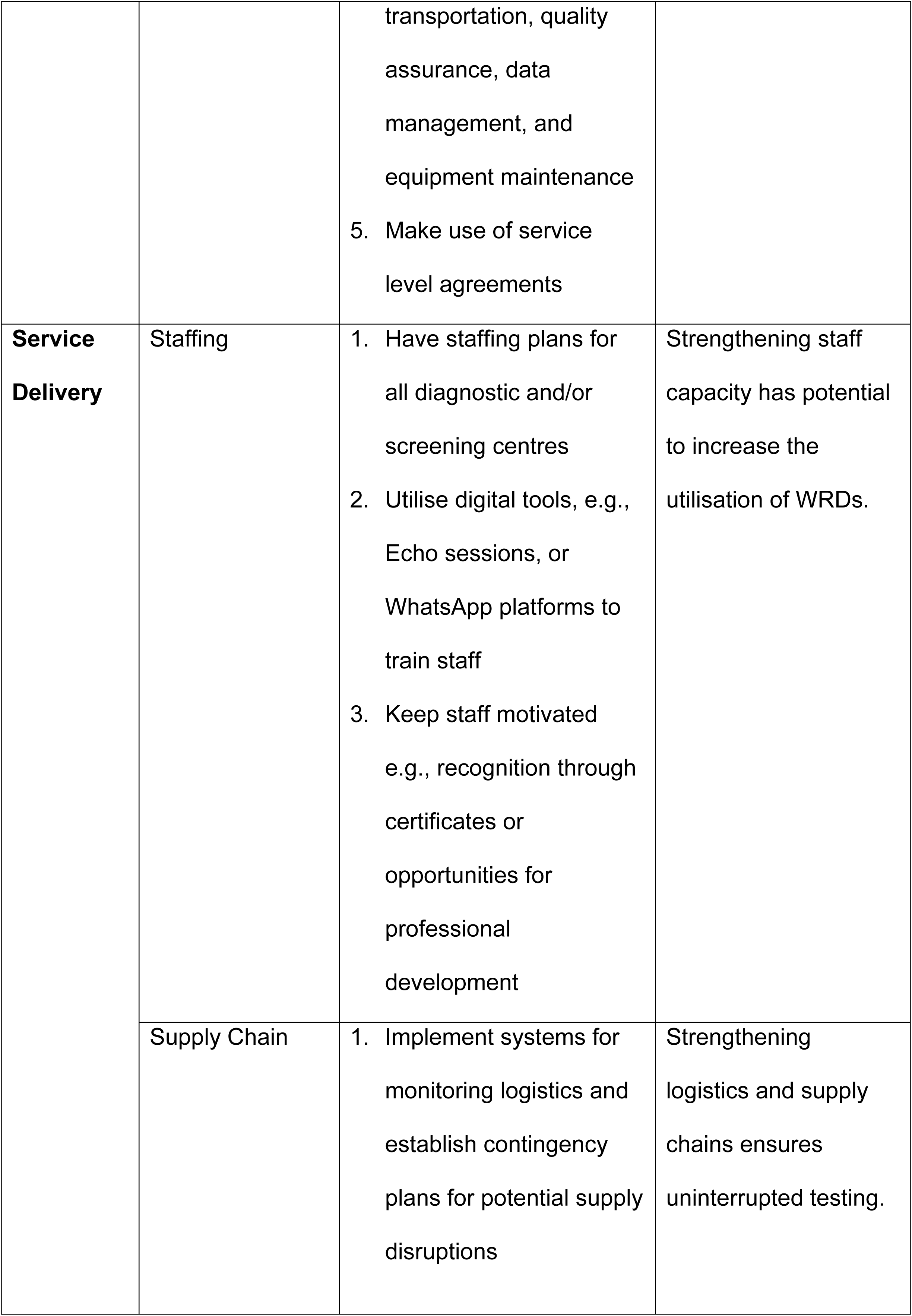
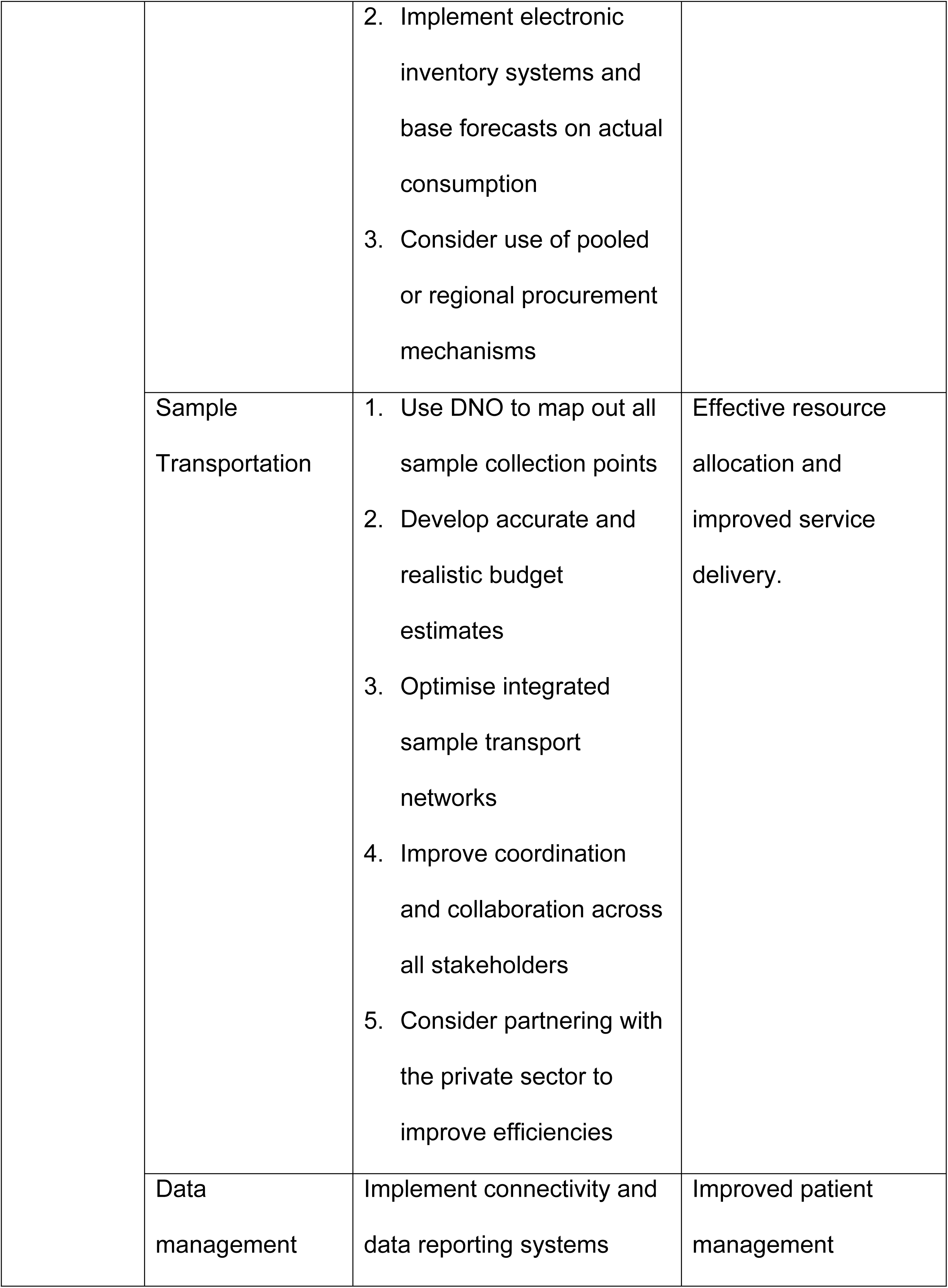
Strategies to reach universal access to WRDs.

| Level | Barrier | Mitigation Strategy | Impact |
| --- | --- | --- | --- |
| **Policy & Strategic Planning** | Financing | 1. Resource reallocation with focus on increasing domestic funding | In the context of reduced donor/external funding, increasing |
|  |  | 2. Inclusion of laboratory directors in technical working groups for planning and funding requests<br>3. Implement annual review of diagnostic network to inform decision-making | domestic funding for diagnostic networks is essential for sustainable WRD use. |
|  | Strategic planning for WRD use | 1. Program commitments and supportive policies for deployment of WRDs<br>2. Use of diagnostic network optimisations for strategic placement of tools<br>3. Use program data in planning, e.g., maintaining updated master facility and equipment lists<br>4. Plan and budget for all aspects of WRD use including infrastructure, staffing, sample<br>transportation, quality assurance, data management, and equipment maintenance<br>5. Make use of service level agreements | Long term planning guided by program data for the use and maintenance of WRDs. |
| **Service Delivery** | Staffing | 1. Have staffing plans for all diagnostic and/or screening centres<br>2. Utilise digital tools, e.g., Echo sessions, or WhatsApp platforms to train staff<br>3. Keep staff motivated e.g., recognition through certificates or opportunities for professional development | Strengthening staff capacity has potential to increase the utilisation of WRDs. |
|  | Supply Chain | 1. Implement systems for monitoring logistics and establish contingency plans for potential supply disruptions | Strengthening logistics and supply chains ensures uninterrupted testing. |
|  |  | 2. Implement electronic inventory systems and base forecasts on actual consumption<br>3. Consider use of pooled or regional procurement mechanisms |  |
|  | Sample Transportation | 1. Use DNO to map out all sample collection points<br>2. Develop accurate and realistic budget estimates<br>3. Optimise integrated sample transport networks<br>4. Improve coordination and collaboration across all stakeholders<br>5. Consider partnering with the private sector to improve efficiencies | Effective resource allocation and improved service delivery. |
|  | Data management | Implement connectivity and data reporting systems | Improved patient management |

### Theme 1: Optimise WRD capacity

All respondents highlighted the existence of policies and strategic plans for the use of WRDs as essential for expanding WRD capacity. This included revision of national guidelines to emphasize the use of WRDs as well as redeployment of devices to increase access. One country reported increasing WRD use by prioritizing equipment placement based on access rather than just utilization, thus ensuring access to areas where sample transportation is challenging. Another country also reported that diversifying the install base of molecular tools, that is Xpert, TB-LAMP and Truenat, had contributed to a higher proportion of individuals with TB being tested using WRDs. Diagnostic network and integrated testing were identified as key enablers for increasing WRD use. However, consumable/cartridge stock outs, limited resources for reagents, and delays with equipment maintenance were key barriers to WRD use in five of the six countries.

> “As much as countries can have equipment capacity, factors like geographical location, sample referral systems, staffing, and equipment downtime will affect the number of presumptive TB that are tested. Additionally, most of the times the equipment is placed in high tier facilities and in the urban areas, and not at PHC level” (TB Diagnostics Officer, NTP)

### Theme 2: Strengthen sample transport and access to testing

Strengthening sample referral networks and intensifying community active case finding were identified as key facilitators for WRD use. Most countries reported that integrated sample transportation had improved turnaround times and enabled sample collection in places that could not previously be reached by the siloed sample transport approach. However, one country reported that difficulties in implementing the shared sample transport systems, citing challenges with funding and coordination across programs and different levels of the health system. Most countries also reported sample transport challenges due to inadequate fuel allocation or insufficient funds for sample transportation and poor road conditions which made certain areas inaccessible, particularly during the rainy season.

> “Inadequate contact tracing, insufficient mobilisation/awareness among the community and service providers to promote screening for all coughers, as well as difficulty in implementing the cough monitoring approach due to lack of funding are barriers for the identification of presumptive TB” (Laboratory Focal Point, NTP)

> “For us the main issue is sample transportation. We have tried to have on demand courier services to cater for presumptive TB wherever they show up at a facility. However, there are some areas where the courier can only go once a month. Certain areas are also cut off most of the year due to flooding so in those cases, microscopy is used. The other challenge we face is that some patients fail to submit sputum, or there are others for whom sputum is never collected, so even though the capacity for testing presumptive TB is there, these patients will be diagnosed clinically” (TB Diagnostics Officer, NTP)

> “Poor road conditions, particularly in rural areas, and traffic congestion in urban areas pose major obstacles. Additionally, the actual cost incurred by the structures far exceeds the planned cost in this integrated sample transport system” (Laboratory Focal Point, NTP)

### Theme 3: Strengthen financing, staffing and infrastructure

Funding was highlighted by all countries as a major barrier to WRD use. Two countries reported that the insufficient number of rapid diagnostic devices was a funding issue, and this also impacted sample transportation, reagents, equipment maintenance and staffing. Additionally, one country reported lack of staff motivation as a major deterrent to the use of molecular tools. Two countries reported that poor infrastructure limited the capacity of peripheral laboratories to conduct molecular testing.

> “Generally, low investment by countries in disease control is a problem. There is need to involve the laboratory in making final decisions regarding funding requests.” (Head of Laboratory, NTRL)

> “For us, a shortage of trained staff is a barrier as some staff are not confident to use molecular tools. However, staff motivation is also a major challenge. The staff say that they use expensive machines but are not well paid and therefore prefer smear microscopy.” (Manager, NTRL)

## Discussion

Despite remarkable advancements in TB diagnostics over the past decade, sputum microscopy remains the mainstay of diagnosis in most high burden TB countries. Our analysis of data from 24 African countries shows that the use of rapid molecular tests remains below the universal testing target, with only 7 countries on track to meet the UNHLM target. In 2023, 61% of notified people with TB in the African countries analysed were tested with a WRD, above the global average of 48%, but below the target of universal access. This reflects critical gaps in implementation, highlighting the need for concerted efforts to accelerate adoption of WRDs, track progress and reinforce accountability.

Theoretically, the WRD target should be the easiest to meet, because the tools are available to ensure universal access. The slow pace of uptake, however, suggests that technological advancements, including simplified test workflows, are not the panacea we hoped for. The country responses showed that the policies are available and have been widely adopted, as has been previously reported in an analysis of 43 high burden TB countries that showed >80% had national policies on WRDs as the initial diagnostic test for all presumptive TB (9). This suggests persisting bottlenecks in translating policies to action. As our results show, some of these barriers include inconsistent implementation of diagnostic algorithms in countries, with only 46% of the countries that submitted benchmark data reporting that a WRD algorithm was available in all facilities. This also frequently comes up as a gap/weakness in countries where we have led diagnostic network assessments or optimisations in Africa. In some countries, initial testing algorithms restricted Xpert testing to high-risk groups, and the transition to an updated algorithm at the subnational or peripheral level has been slow and inconsistent, leading to low utilisation of instruments.

Many of the diagnostic network constraints reported by the country respondents relate to infrastructure, staffing, and funding, in agreement with previous publications (10, 11). The responses showed that underutilization of WRDs is due to infrastructural challenges, lack of trained staff and maintenance challenges. Unstable electricity, inadequate operational standards and low testing demand have also been previously reported as barriers to the use of GeneXpert devices (12). A surprising barrier reported by one of the countries was the low staff motivation to use WRDs and preference for microscopy testing, in contrast to the high acceptability of molecular tools previously reported in multiple African countries (13, 14). Continuous training, recognition or performance-based rewards might help with increasing motivation in that setting.

All respondents identified funding as a key barrier. That diagnostics, across multiple diseases, not just TB, are underfunded is well established (15). Less than 3% of the $5.3 billion spent annually on global TB control is on diagnostics (8). However, considering current funding cuts, the situation becomes particularly dire. Over half the $898 million mobilized for TB prevention, diagnosis and treatment across the African continent in 2023 was from international funding (8). Recent surveys that we and others have conducted show that the impact of these cuts on the diagnostic networks is primarily on staffing, sample transport networks, equipment maintenance and quality assurance and control systems (16, 17). Additionally, in our survey of 16 African laboratory directors, several countries also reported reduced access to reagents and supplies and reduced scope of diagnostic testing (17). Concerningly, more than 80% reported they would not be able to sustain operations beyond 12 months if the funding cuts persist. The fact that the impact of these cuts is on the same areas identified as challenges to increasing access to WRDs, highlights a critical need for programs to secure sustainable funding, advocate for increased domestic financing and strengthen partnerships with the private sector. It is imperative, now more than ever, to implement the principles of the Lusaka Agenda (18), and ensure that diagnostics are embedded within a universal health coverage framework and aligned with domestic financing plans. This will necessitate a shift towards a primary healthcare model, and that may well be possible with new near point of care assays on the horizon.

Stock outs of TB test commodities and medications are a recurrent theme in most African countries, with low stock accuracy, delayed reporting and poor communication as main hurdles (19, 20). A review of the TB commodities supply chain in the WHO African region showed that countries that relied on domestic mechanisms for procurement were more vulnerable to stock outs due to inadequate funding and longer procurement lead times (21). Pooled procurement mechanisms to achieve volume concession for tests could assist with reducing the prices. This was recently demonstrated in the price reductions for molecular tests and treatment regimens for drug-resistant TB (22, 23). Global or regional procurement may also be beneficial for countries with smaller volumes to access concessional pricing for service and maintenance. However, lower prices alone without strengthening the health system will not necessarily translate into more people being tested and treated.

Continuous network optimization is therefore critical to enable programs to identify and promptly address challenges. For example, diagnostic network optimisation/assessments can assist countries to assess their capacity to test all people with TB, tailor the network, and inform operational planning to ensure equitable access to diagnostics (24, 25). These optimisations also present an opportunity for countries to assess and enhance efficiency of integrated laboratory networks, thus offering a cost-effective option for maximising population coverage across multiple diseases (26). To support countries in monitoring their progress towards achieving the UNHLM WRD target, we have developed a TB-WRD scorecard aligned to the WHO Diagnostic Standard. This scorecard builds on ASLM’s laboratory network assessment (LABNET) model and seeks to assist countries to use and interpret existing TB data to identify the gaps within the TB diagnostic cascade, plan and allocate resources effectively for targeted interventions. It is currently being piloted in the 24 LabCoP countries and results will be discussed at our annual meeting.

A key strength of our analysis was combining data submitted to the WHO with survey responses from national TB program and TB reference laboratory staff to identify contextual factors influencing WRD use. We also purposively selected respondents to reflect different geographical regions and levels of WRD use, enriching the responses and capturing a wide spectrum of opinions. However, we only reached out to the selected countries and not all 24 countries within the LabCoP network which may limit generalisability of the findings. A planned report from our annual meeting will capture responses from all 24 countries. Furthermore, we did not triangulate the survey findings with other data sources such as national strategic plans or diagnostic network reports which might introduce bias. Nonetheless, the themes that emerged as barriers align with previous reports suggesting these are key considerations for implementation of diagnostic tools.

## Conclusion

Despite remarkable advancements in TB diagnostics, our analysis shows that the use of rapid molecular tests is inconsistent and remains below the target of 100% WRD testing, with only 7 of 24 countries on track to achieve it. With only 5 years left to the realization of the 2030 Agenda for Sustainable Development, there is need for accelerated action to meet the targets of eliminating the TB epidemic. While universal access to quality TB diagnosis is central to the first pillar of the End TB Strategy, the slow pace of adoption of rapid molecular diagnostic tools suggests that diagnostics alone cannot overcome systemic barriers. Persistent challenges in infrastructure, funding, staffing and sample transportation require a data driven, systems-level approach that holistically evaluates the gaps in the diagnostic network and implements targeted interventions focused on improving the entire TB cascade.

## Data Availability

All data supporting our findings are contained within this manuscript. Datasets used for the quantitative analysis are open access and available from the WHO Global TB website. This is an open access article distributed under the terms of the Creative Commons Attribution License, which permits unrestricted use, distribution, and reproduction in any medium, provided the original author and source are credited.

## Notes

### Competing Interest Statement

The authors have declared no competing interest.

### Funding Statement

This work was supported by the Gates Foundation (INV-038883). The content of this paper is solely the responsibility of the authors and does not necessarily represent the official views of the Gates Foundation.

## References

1. WHO. WHO consolidated guidelines on tuberculosis: module 3: diagnosis 2025 [Available from: https://www.who.int/publications/i/item/9789240107984.

2. Subbaraman R, Nathavitharana RR, Mayer KH, Satyanarayana S, Chadha VK, Arinaminpathy N, et al. Constructing care cascades for active tuberculosis: A strategy for program monitoring and identifying gaps in quality of care. PLOS Medicine. 2019;16(2):e1002754.

3. WHO. Global TB Report 2024 [Available from: https://www.who.int/teams/global-programme-on-tuberculosis-and-lung-health/tb-reports/global-tuberculosis-report-2024.

4. WHO. Political declaration of the UN General-Assembly High-Level Meeting on the Fight Against Tuberculosis 2018 [Available from: https://www.who.int/publications/m/item/political-declaration-of-the-un-general-assembly-high-level-meeting-on-the-fight-against-tuberculosis.

5. WHO. Status Update: Reaching the targets in the political declaration of the United Nations general assembly high level meeting on the fight against tuberculosis 2023 [Available from: https://cdn.who.int/media/docs/default-source/un-high-level-meeting-on-tb/who-ucn-tb-2023.4.pdf.

6. WHO. The second United Nations high-level meeting on TB: new global pledge to end the TB epidemic 2023 [Available from: https://www.who.int/teams/global-tuberculosis-programme/tb-reports/global-tuberculosis-report-2023/featured-topics/un-declaration-on-tb.

7. WHO. WHO standard: universal access to rapid tuberculosis diagnostics 2023 [Available from: https://www.who.int/publications/i/item/9789240071315.

8. WHO. Global TB Report Data 2024 [Available from: https://www.who.int/teams/global-programme-on-tuberculosis-and-lung-health/data.

9. Stop TB Partnership. Step Up for TB 2024 [Available from: https://www.stoptb.org/what-we-do/advocate-endtb/step-tb-national-tb-policies.

10. Ntinginya NE, Kuchaka D, Orina F, Mwebaza I, Liyoyo A, Miheso B, et al. Unlocking the health system barriers to maximise the uptake and utilisation of molecular diagnostics in low-income and middle-income country setting. BMJ Glob Health. 2021;6(8).

11. Piatek AS, Wells WA, Shen KC, Colvin CE. Realizing the “40 by 2022” Commitment from the United Nations High-Level Meeting on the Fight to End Tuberculosis: What Will It Take to Meet Rapid Diagnostic Testing Needs? Global Health: Science and Practice. 2019:GHSP-D-19-00244.

12. Dörfler J, Ravololohanitra OG, Decroo T, Franke MA, Emmrich J, Pasteur GI, et al. Underutilisation of GeneXpert devices for TB diagnosis: a missed opportunity. IJTLD Open. 2025;2(7):440–2.

13. Engel N, Ochodo EA, Karanja PW, Schmidt BM, Janssen R, Steingart KR, et al. Rapid molecular tests for tuberculosis and tuberculosis drug resistance: a qualitative evidence synthesis of recipient and provider views. Cochrane Database of Systematic Reviews. 2022(4).

14. Nwokoye N, Ihesie A, Olabamiji J, Ochei K, Eneogu R, Umoren M, et al. Exploring the perspectives of healthcare workers and Program managers on the use of Truenat as a new tool for TB and DR-TB diagnosis in Nigeria: A qualitative study. PLOS ONE. 2025;19(12):e0316204.

15. Fleming KA, Horton S, Wilson ML, Atun R, DeStigter K, Flanigan J, et al. The Lancet Commission on diagnostics: transforming access to diagnostics. The Lancet. 2021;398(10315):1997–2050.

16. Stop TB Partnership. Report in the impact of US government funding halt on TB responses in high TB burden countries 2025 [Available from: https://www.stoptb.org/sites/default/files/documents/Disruption%20US%20FUNDING%20halt030325.pdf.

17. ASLM. The impact of a temporary suspension of United States Government (USG) funding on laboratory services in African partner countries 2025 [Available from: https://aslm.org/wp-content/uploads/2025/05/Impact-Report-USG-.pdf.

18. FGHI. The Lusaka Agenda: Conclusions of the Future of Global Health Initiatives process 2023 [Available from: https://futureofghis.org/final-outputs/lusaka-agenda/.

19. Tiye K, Gudeta T. Logistics management information system performance for program drugs in public health facilities of East Wollega Zone, Oromia regional state, Ethiopia. BMC Medical Informatics and Decision Making. 2018;18(1):133.

20. Desale A, Taye B, Belay G, Nigatu A. Assessment of laboratory logistics management information system practice for HIV/AIDS and tuberculosis laboratory commodities in selected public health facilities in Addis Ababa, Ethiopia. Pan Afr Med J. 2013;15:46.

21. Mhazo AT, Miyango S, Palani L, Maponga CC. Tuberculosis commodities supply chain performance in the WHO African region: A scoping review. PLOS Glob Public Health. 2024;4(5):e0003219.

22. Holt E. Price reductions in drug-resistant tuberculosis treatments. The Lancet Microbe. 2025.

23. The Global Fund. Briefing Note: New Pricing for Cepheid GeneXpert Tuberculosis Testing 2023 [Available from: https://www.theglobalfund.org/media/13442/operational_2023-10-cepheid-genexpert-tb-testing_briefingnote_en.pdf.

24. Albert H, Rupani S, Masini E, Ogoro J, Kamene M, Geocaniga-Gaviola D, et al. Optimizing diagnostic networks to increase patient access to TB diagnostic services: Development of the diagnostic network optimization (DNO) approach and learnings from its application in Kenya, India and the Philippines. PLoS One. 2023;18(11):e0279677.

25. Piatek AS. Assessment of TB Diagnostic Networks: A new tool 2018 [Available from: https://www.teachepi.org/wp-content/uploads/Courses/ADV-2018/APiatek_adv_TB_diag2018June20.pdf.

26. Ondoa P, Ndlovu N, Keita MS, Massinga-Loembe M, Kebede Y, Odhiambo C, et al. Preparing national tiered laboratory systems and networks to advance diagnostics in Africa and meet the continent’s health agenda: Insights into priority areas for improvement. Afr J Lab Med. 2020;9(2):1103.

